# Whole-brain computational modeling reveals disruption of microscale brain dynamics in HIV infected individuals

**DOI:** 10.1101/2020.04.29.20084558

**Authors:** Yuchuan Zhuang, Zhengwu Zhang, Madalina Tivarus, Xing Qiu, Jianhui Zhong, Giovanni Schifitto

## Abstract

In this study, we adopted the relaxed mean-field dynamic modeling to investigate structural and functional connectivity in forty-two HIV-infected subjects before and after 12-week of combination antiretroviral therapy (cART) and compared them with forty-six age-matched healthy subjects. Microscale brain dynamics were modeled by a set of parameters including two region-specific microscale brain properties, recurrent connection strengths, and subcortical inputs. We also analyzed the relationship between the model parameters (i.e. the recurrent connection and subcortical inputs) and functional network topological characterizations. The results show that untreated HIV-infected individuals have disrupted local brain dynamics that in part correlate with network topological measurements. Notably, after 12 weeks of cART, both the microscale brain dynamics and the network topological measurements improved and were closer to those in the healthy brain. This was also associated with improved cognitive performance, suggesting that improvement in local brain dynamics translates into clinical improvement.

## 1. Introduction

There were approximately 37.9 million people globally living with the HIV in 2018 according to world health organization (WHO, https://www.who.int/gho/hiv/en/). Since the introduction of antiretroviral therapy (ART), there has been a significant decrease in the mortality rate of people infected with HIV, and a dramatic decrease in the incidence of HIV-associated dementia (HAD) (Saylor et al., 2016). However, the prevalence of HIV-associated neurocognitive disorders (HAND) is increasing (Nabha, Duong, & Timpone, 2013). The cause of the high prevalence of HAND remains unclear. It is likely multifactorial, including early injury prior to starting antiretroviral drugs (HIV infects the brain shortly after transmission), chronic mild neuroinflammation and possibly neurotoxicity of antiretroviral drugs (Chang & Shukla, 2018; Fois & Brew, 2015).

MRI-based neuroimaging techniques are valuable in the investigation of HIV-infection associated neuropathology (Chang & Shukla, 2018). HIV-infected subjects have reduced cortical thickness and subcortical brain volumes (Chang & Shukla, 2018; Sanford, Fellows, Ances, & Collins, 2018), and reduced functional connectivity (FC) (Samboju, Philippi, Chan, Cobigo, Fletcher, Robb, Hellmuth, Benjapornpong, Dumrongpisutikul, Pothisri, Paul, Ananworanich, Spudich, Valcour, Search, et al., 2018; Zhuang et al., 2017). Both imaging metrics have been associated with impaired cognitive performance when compared with healthy controls (Samboju, Philippi, Chan, Cobigo, Fletcher, Robb, Hellmuth, Benjapornpong, Dumrongpisutikul, Pothisri, Paul, Ananworanich, Spudich, Valcour, Search, et al., 2018; Sanford et al., 2018; Zhuang et al., 2017). Brain injury at a microstructural level can be quantified by diffusion tensor imaging (DTI) metrics, such as decreased fractional anisotropy (FA) and increased mean diffusivity (MD). Both FA and MD abnormalities have been previously reported in HIV-infected subjects (Stebbins et al., 2007; Underwood et al., 2017; Zhu et al., 2013). Graph theoretical analysis of structural and functional connectome has also been used to show brain network topological changes in HIV-infection (Abidin et al., 2018; Bell et al., 2018b; Chockanathan, AM, Abidin, Schifitto, & Wismuller, 2018; Thomas, Brier, Ortega, Benzinger, & Ances, 2015).

Overall, there is evidence that brain injury is present and measurable via MRI in HIV infected individuals. However, most of the previous studies have investigated one modality at a time, whereas the integration of multi-modalities in HIV-related studies has not been fully explored yet. Large-scale whole-brain dynamic modeling is a promising approach to further quantify the integrated contribution of structural and functional connectivity in CNS injury. This approach allows simulating resting-state fluctuations emerging from the interaction between brain regions, constrained by the anatomical connections derived from DTI, and effectively integrates functional connectivity (FC) and structural connectivity (SC) (Honey et al., 2009; Mollink et al., 2019; Surampudi et al., 2019; P. Wang et al., 2019). Furthermore, this approach has already provided some insights in connectome disruption in other neurological disorders such as Alzheimer’s disease and Parkinson’s disease (Alderson et al., 2018; Deco & Kringelbach, 2014; Demirtas et al., 2017; Jirsa, Sporns, Breakspear, Deco, & McIntosh, 2010; Proix, Bartolomei, Guye, & Jirsa, 2017). A novel whole-brain modeling technique, named relaxed mean field dynamic modeling (rMFM) (P. Wang et al., 2019) has been proposed to simulate local brain dynamics. Previous whole-brain modeling studies assumed that local microscale properties, the recurrent connection strengths and subcortical inputs, were the same across entire brain, while the rMFM modeling method relaxed these two parameters to be heterogeneous across different brain regions. Two microscale brain properties, recurrent connection strengths and subcortical inputs, can be derived from this generative brain dynamic modeling by tuning the model to fit the simulated FC to empirical FC.

In this study, we applied the rMFM (P. Wang et al., 2019) to assess CNS changes in a cohort of HIV infected treatment-naïve patients at baseline (HIV+BSL) and after 12 weeks of cART treatment (HIV+12wk), and compared them to healthy controls (HC). The overreaching goal was to investigate potentially new imaging biomarkers that could be sensitive to CNS changes and thus helpful in monitoring CNS disease progression and response to treatment. Firstly, we built the rMFM whole-brain dynamic models for the three groups (HC, HIV+BSL, HIV+12wk) respectively. We then compared the microscale brain properties (the recurrent connection strength and subcortical inputs) across the three groups. This was followed by investigating the topological changes among the three groups using traditional graph theoretical analysis from both global and regional perspectives. Subsequently, we investigated the association of nodal graph theoretical measurements with microscale brain properties and neuropsychological tests scores.

## 2. Results

### 2.1. Demographics

Forty-two HIV+ subjects were age matched with forty-six HC. The HIV+ subjects were scanned before starting the cART treatment, and scanned, on average, after 12-week of the initiation of cART treatment. At baseline, twenty-one HIV+ individuals had normal cognitive performance, twenty had Asymptomatic Neurocognitive Impairment (ANI), and one had Mild Neurocognitive Disorder (MND). The overall cognitive performance, based on the summary Z score of all cognitive tests, was significantly higher in the HC compared to HIV infected individuals. The mean CD4 count and HIV RNA plasma levels at baseline were 515.8±42.3 cells/mm^3^, and 4.254±0.164 log_10_ copies/ml, respectively. After 12 weeks, mean CD4 cell count and HIV RNA levels were 566.4±44.5 cells/mm^3^ and 2.675±2.641 log_10_ copies/mL, respectively.

### 2.2. FC simulation using rMFM

We performed rMFM simulations with 25 different random initializations for each cohort respectively, 75 simulations in total. Supplementary Fig. 1A shows the increase of similarity within 500 iterations for model optimization. The similarities within each cohort are consistent, and all above 0.55, indicating that the rMFM whole-brain dynamic model yields good simulation of FC from SC in the training dataset of each cohort. The similarity between simulated FC and empirical FC shows high consistency across different random initializations.

We found that using different initialization parameters had little effect on the final similarity results (see Supplementary Fig. 1B). The variance of the maximum similarity Z-score for 25 random initializations for HC, HIV+BSL, and HIV+12wk are 5.438 × 10^−5^, 8.886 × 10^−5^, and 4.210 × 10^−5^, respectively. Different initialization parameters also had little effect on the final rMFM model parameters (see Supplementary Fig. 2), including the recurrent connection w, excitatory subcortical input I, global scaling of the structural connectivity G, and neuronal noise *σ*. Therefore, to minimize this effect, for each cohort, the optimum rMFM model was adopted from the averaged values across the five runs which had the highest similarities.

### 2.3. Model validation

We validated the rMFM model parameters for each cohort using the test dataset. Then we calculated the simulated neuronal activity for each brain region by feeding the empirical SC from the test dataset into the fitted rMFM model for each cohort. We calculated the simulated BOLD signal for each region by feeding the simulated neuronal activity to the Balloon-

Windkessel hemodynamic model. The simulated FC was calculated by Pearson’s correlation. We run 1000 simulations with different random initializations for each cohort using the test dataset. The mean simulated FC matrices for each cohort was calculated as the average across the 1000 simulations (see Fig. 1B, 1E, 1H). The correlation between the averaged simulated and empirical FC in the test dataset are 0.498, 0.442, and 0.525 for HC, HIV+BSL, and HIV+12wk, respectively (see Fig. 1C, 1F, 1I). The empirical SC-FC correlation coefficients in the test dataset are 0.401, 0.409, and 0.414, respectively, suggesting that the use of rMFM model for simulating FC from SC improved their agreements by 24.2%, 8.1%, and 26.8%.

**Fig. 1.**
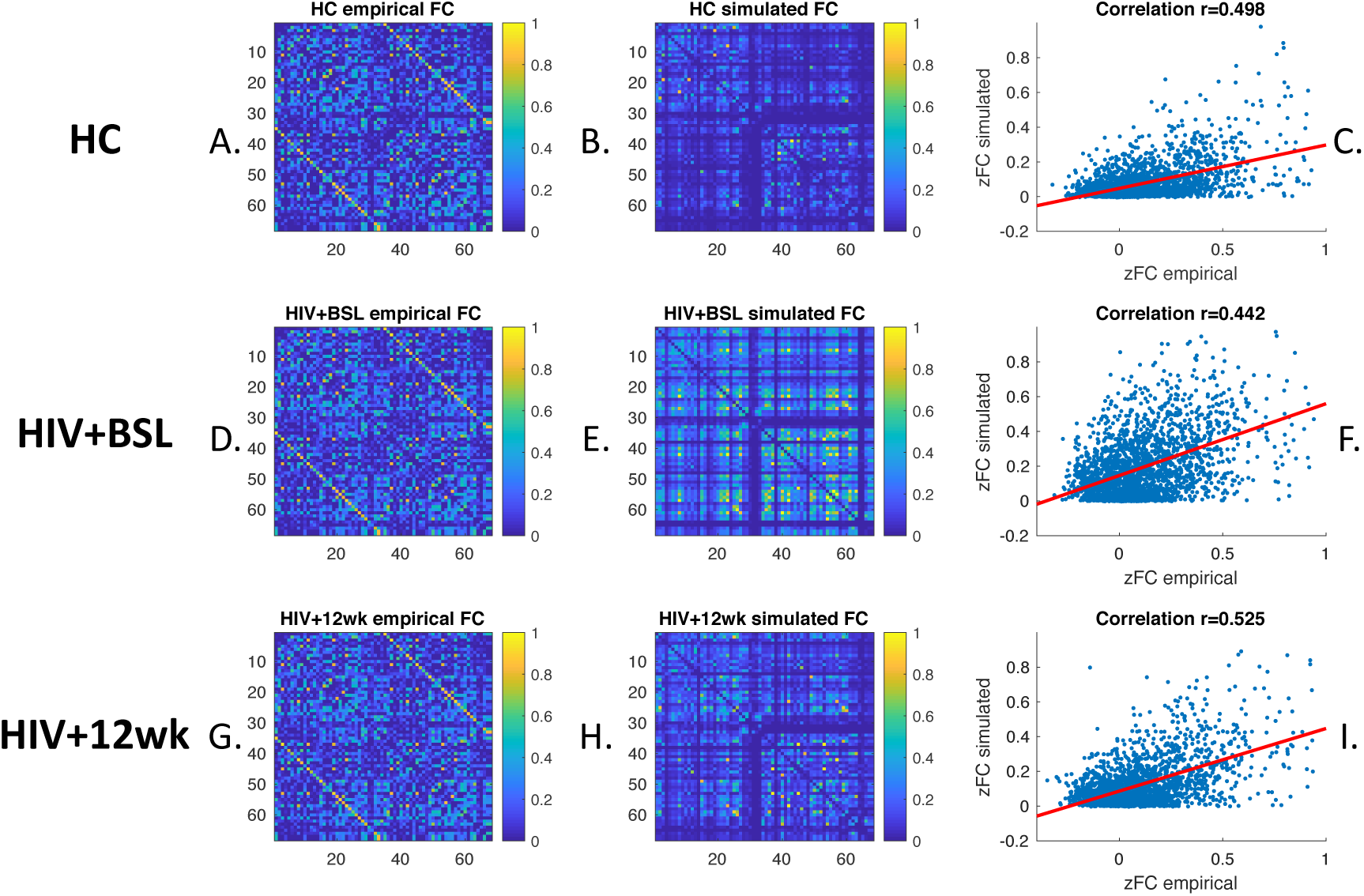
rMFM simulated FC. The empirical and rMFM models generated the simulated FC for HC, HIV+BSL, and HIV+12wk, respectively. Left column A) D) G): the averaged empirical FC in test dataset for each cohort. Middle column B) E) H): the averaged simulated FC. Right column C) F) I): The correlation between empirical and simulated FC.

Further validation steps including: 1) using a different training/test dataset, 2) increase to 1000 iterations, and 3) reproducing results using Destriuex atlas, which can be found in the supplementary materials.

### 2.4. rMFM model parameters comparison between HIV and HC

The microscale brain dynamic properties of the three groups were investigated by comparing the rMFM model parameters, w and I. Here we compared these two regional values between cohorts to identify which brain regions changed significantly with a treatment intervention. The recurrent connection strength w is shown in Fig. 2, while the subcortical inputs I are shown in Fig. 3.

**Fig. 2.**
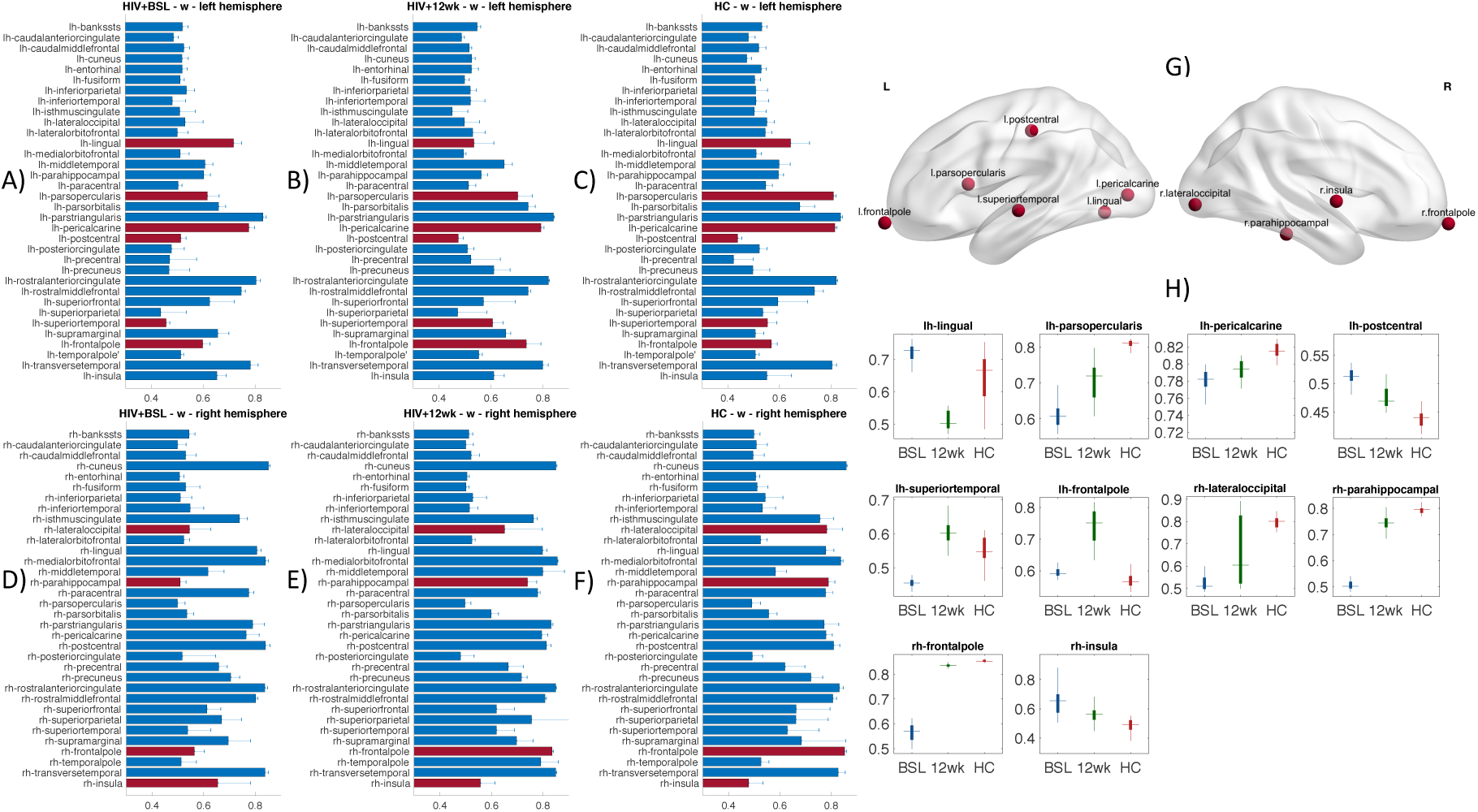
Recurrent connection strengths. A-F show the regional recurrent connection strength (*w_i_*) in three groups; A, D) HIV+BSL, B, E) HIV+12wk, C F) HC. A, B, C – left hemisphere; D, E, F – right hemisphere. The bars and error bars indicate the mean and standard deviation of the recurrent connection strength for each brain region, the red bars indicate the regions which are significantly different between cohorts (FDR p < .01). G). the regional recurrent connection changes in these nodes (red bars in Fig. A-F) plotted on a smoothed brain surface indicate their anatomical locations. H). bar plots show the recurrent connection strength for each ROIs. Red: HC, Blue: HIV+BSL, Green: HIV+12wk.

**Fig. 3.**
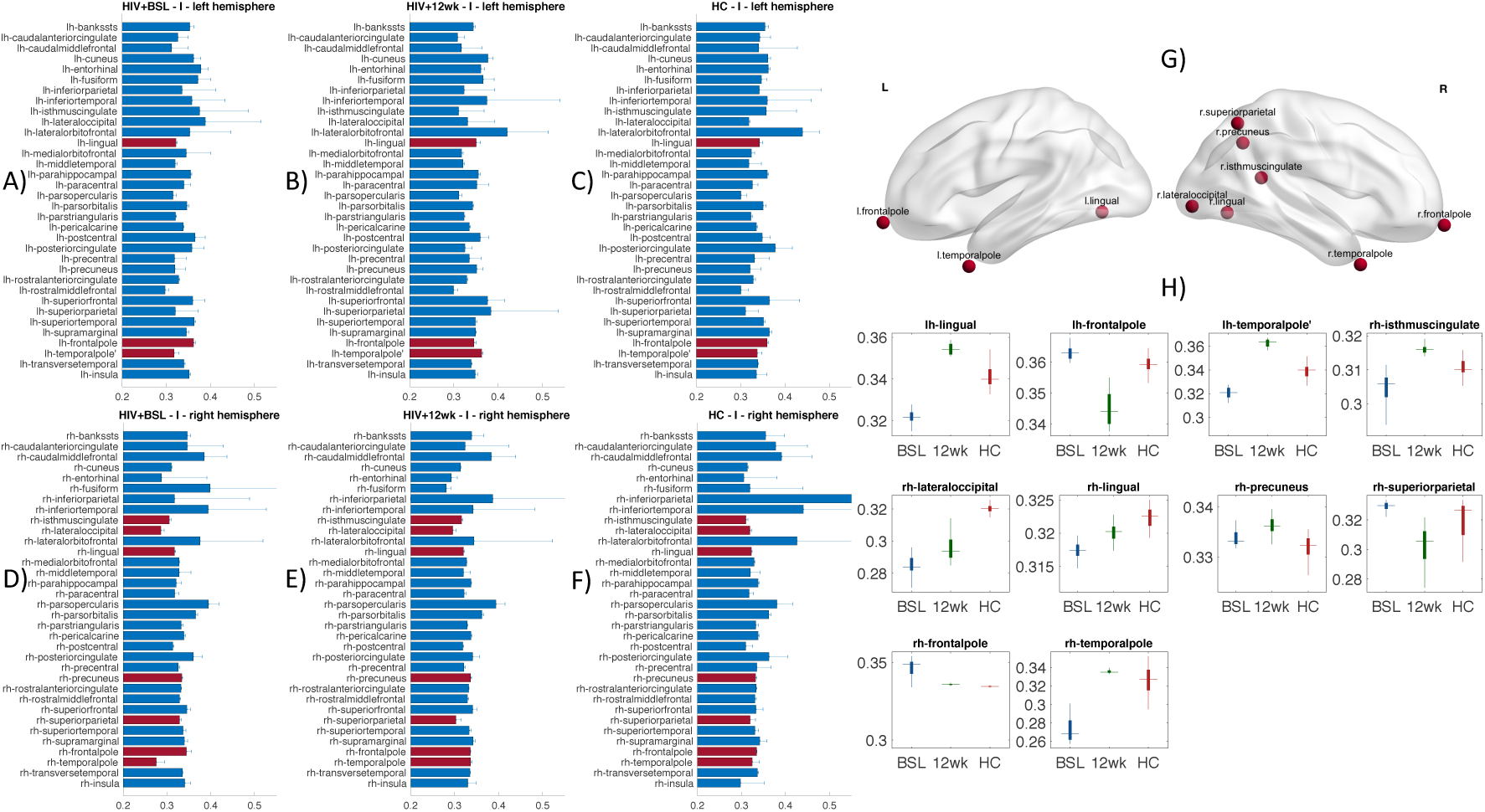
Subcortical input strengths. A-F shows the subcortical input strength (*I_i_*) in the three groups: A, D) HIV+BSL, B,E) HIV+12wk, C, F) HC, in both hemisphere; A, B, C – left hemisphere; D, E, F – right hemisphere. The bars and error bars indicate the mean and standard deviation of the subcortical input strength for each brain region, the red bars indicate the regions which are significantly different between cohort (FDR p < .01). G). the subcortical inputs changes in these nodes plotted on a smoothed brain surface indicate their anatomical locations. H). bar plots show the subcortical input strength for each ROI. Red: HC, Blue: HIV+BSL, Green: HIV+12wk.

The recurrent connection strengths for each region and cohort w_i_, are reported in Fig. 2A-2F. There were ten regions that were significantly different when comparing HIV+BSL, HIV+12wk, and HC (shown in Fig. 2 red bar, also shown in Fig. 2G, FDR corrected p<0.01). These regions are the left frontal pole, opercular part of inferior frontal gyrus (pars opercularis), superior temporal gyrus, postcentral gyrus, pericalcarine cortex, lingual gyrus; right frontal pole, insula, parahippocampal gyrus, and lateral occipital lobe. Their anatomical locations are shown in Fig. 2G. The boxplots in Fig. 2H show the recurrent connection strength w in these regions, there are nine out of ten regions showing clear transitions toward healthy controls values after twelve weeks of cART treatment. The highest differences of recurrent connections between HC and HIV+BSL were found in right frontal pole, right lateral occipital lobe, right parahippocampal gyrus, right insula, right supramarginal gyrus, and left supramarginal gyrus (see Supplementary Fig. 4.).

The subcortical input strength for each region I_i_, and for each cohort are reported in Fig. 3A-3F. The red bars indicate the regions with significant differences among the three groups. The regions include the left frontal pole, temporal pole, and lingual gyrus; right frontal pole, temporal pole, superior parietal lobule, precuneus, isthmus of the cingulate gyrus, lateral occipital lobe, and lingual gyrus. Their anatomical locations are shown in Fig. 3G. We also observed that nine out of ten brain regions showed transitions toward HC after twelve weeks of treatment, for example in left and right frontal pole and right precuneus (see Fig. 3H). We also observed that the regions which differed in subcortical inputs I were asymmetric (see Fig. 3G), with more representation in the right posterior regions. The highest differences of subcortical input strength between HC and HIV+BSL were found in right inferio parietal lobule, left inferio parietal lobule, right fusiform, right lateral orbitofrontal, left isthmus of the cingulate gyrus, and left superioparietal lobule (see Supplementary Fig. 3.). These results are also replicated when using Destrieux atlas (see Supplementary Fig. 11.).

### 2.5. Graph theoretical analysis on empirical FC, correlation with recurrent connection strength w

We have investigated the functional network topology using the conventional graph theoretical analysis. We calculated both global and nodal clustering coefficient, network efficiency, shortest path length, and smallworldness. The global network topology differences between groups are reported in Fig. 4. Using the empirical FC, we have found that, the smallworldness (Fig. 4.A), normalized clustering coefficient (gamma) (Fig. 4.B), and global efficiency (Fig. 4.D) were significantly reduced in HIV+BSL compared to HC, in a majority of network sparsity range (sparsity range: 0.05–0.5, FDR corrected p < 0.05). There were no significant differences between HIV+12wk and HC, or HIV+BSL and HIV+12wk after FDR correction. Here, we only report the results for the binarized FC matrix across different network sparsity. Similar results were reproduced using the weighted FC matrix (Supplementary Fig. 12). We also investigated the network topology on the empirical SC but didn’t find any statistically significant difference between groups.

**Fig. 4.**
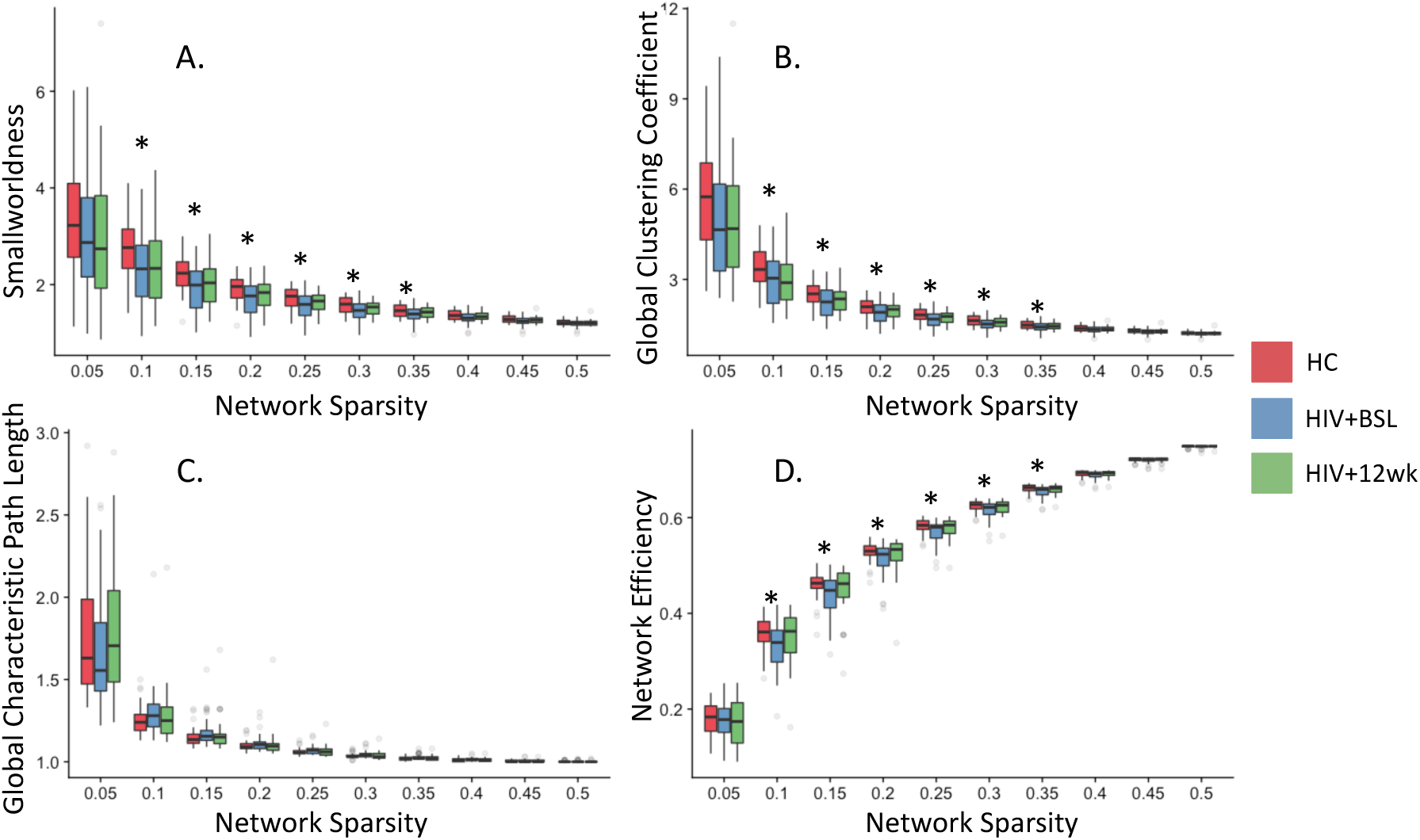
Graph theoretical measurements. Graph theoretical measurements on binarized functional connectivity matrix across network sparsity range from 0.05 to 0.5. A), smallworldness. B). global clustering coefficient. C). global characteristic path length. D). global network efficiency. * FDR corrected p<0.05. Red: HC, Blue: HIV+BSL, Green: HIV+12wk.

Next we investigated the association between local topological measurements and rMFM model parameters. The nodal clustering coefficient and local efficiency results were correlated with recurrent connection strengths w and subcortical inputs I, as shown in Fig. 5. We found significant correlations between recurrent connection strengths w, nodal clustering coefficient (r = 0.248, p = 0.041), and local efficiency (r = 0.253, p = 0.037) only in the HC group. These relationships were confirmed using Destriuex atlas (Supplementary Fig. 13).

**Fig. 5.**
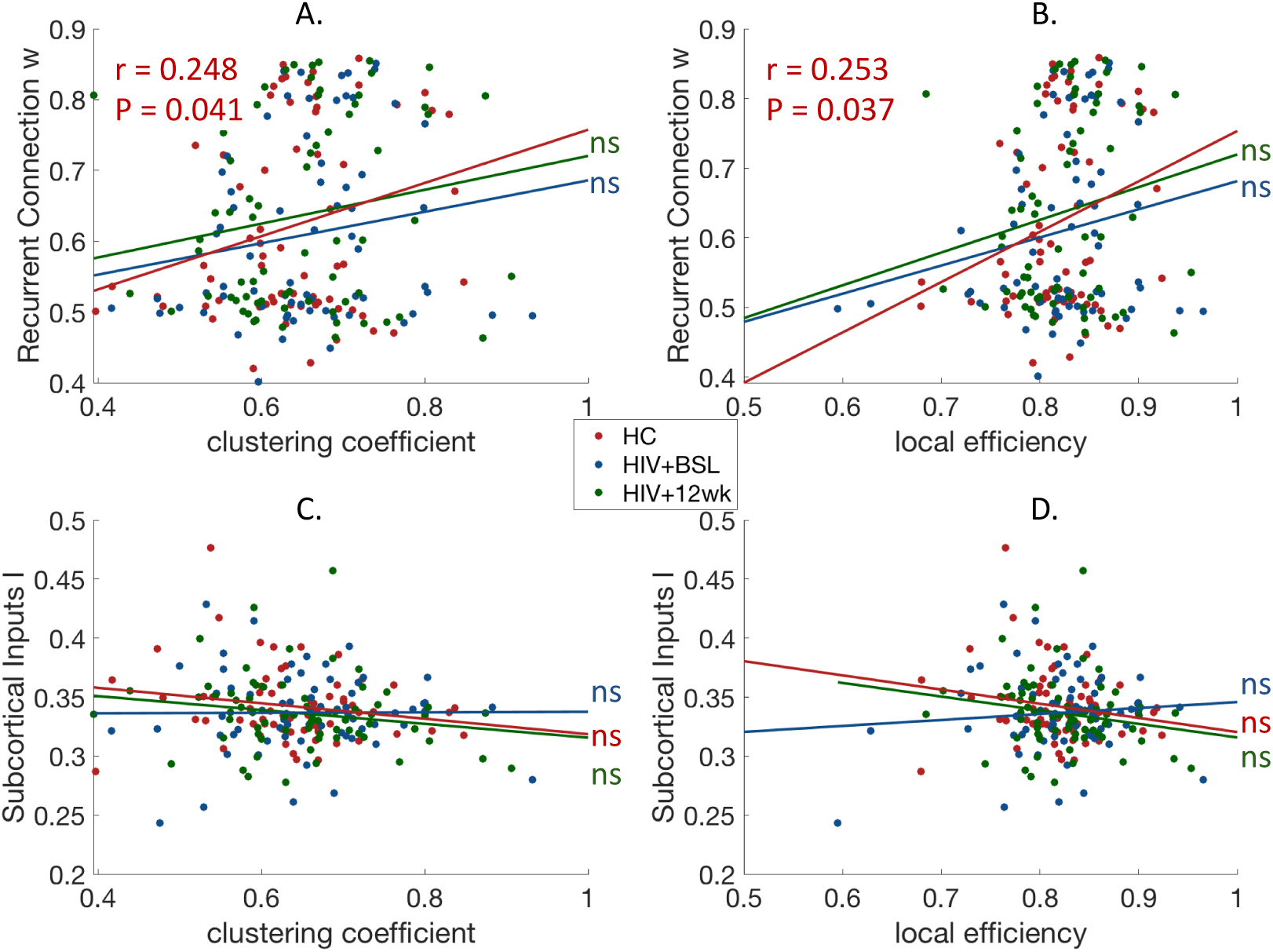
Recurrent connection correlated with network topology. The recurrent connection *w* and subcortical inputs *I* correlated with empirical local network topology. (A) Association between clustering coefficient and recurrent connection strength *w*. (B) Association between local efficiency and recurrent connection strength *w*. (C) Association between clustering coefficient and subcortical inputs *I*. (D) Association between local efficiency and subcortical inputs *I*. Red: HC, Blue: HIV+BSL, Green: HIV+12wk. ns: not significant

### 2.6. Neuropsychological test score comparison, association with FC graph theoretical measurements

As shown in Fig. 6, HIV+BSL subjects had lower overall composite Z-score on the neurophysiologic test battery, and lower motor function score when compared to HC (uncorrected p < .05). However, the HIV+12wk shows no significant difference when compared to HC or to HIV+BSL. Cognitive performance improved in the HIV-infected subjects after 12weeks, as the boxplots show. The neuropsychological test scores in other domains, including speed, attention, learning, memory, executive function, and verbal fluency, although not significantly different between groups, were trending toward better performance in the HC which explain why the summary Z score was significantly different between HIV+BSL and HC. Neuropsychological test results for other cognitive domains that are not significantly different are reported in supplementary materials (Supplementary Fig. 6).

**Fig. 6.**
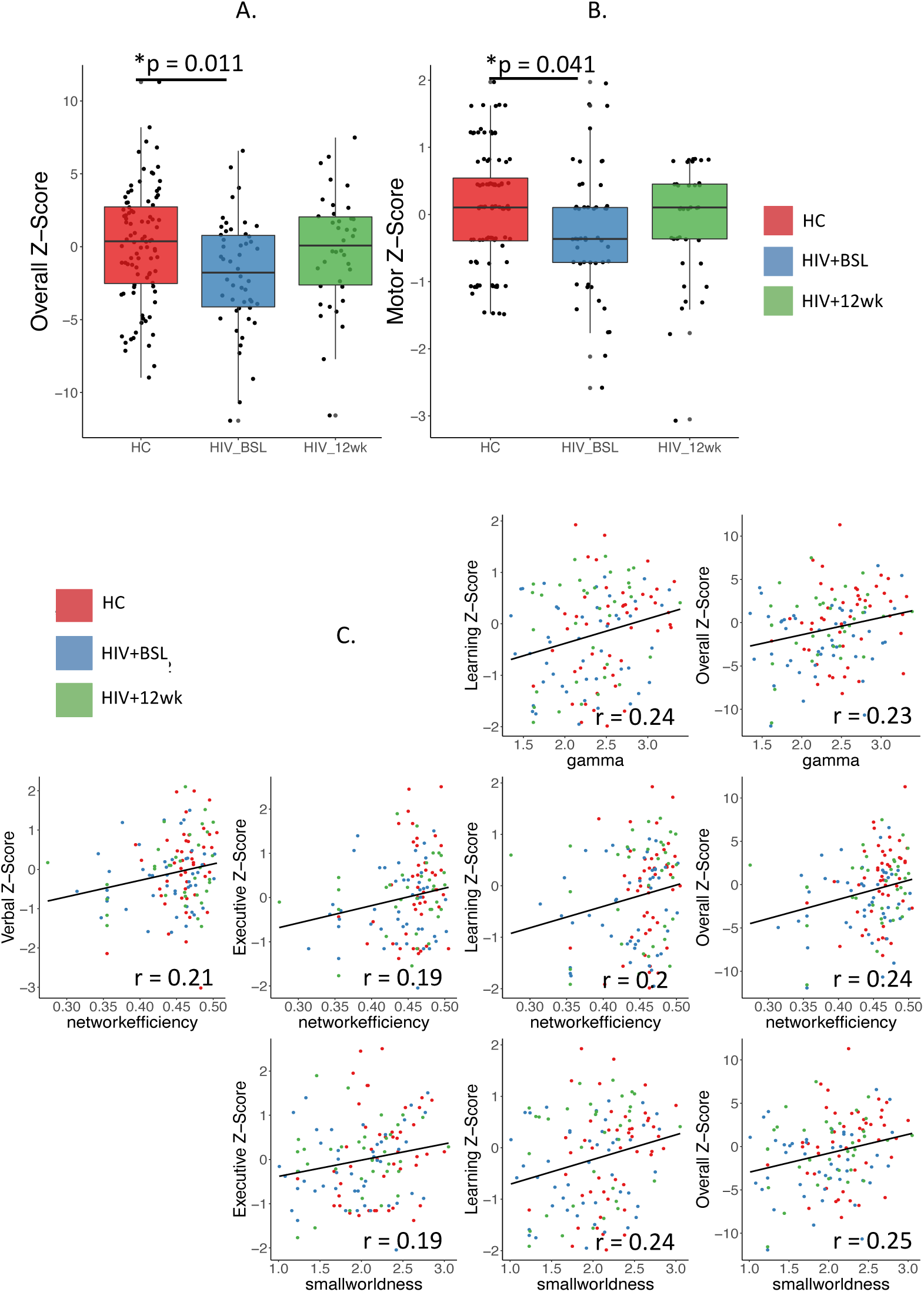
Neuropsychologic test. A) Overall composite Z-score. B: Motor Z-score. C) Linear correlation between empirical FC graph theoretical measurements and Neuropsychological test Z-score. Red: HC, Blue: HIV+BSL, Red: HIV+12wk. All of these plots are reach to significance level: uncorrected p < .05. gamma = normalized network clustering coefficient

We also investigated the relationship between Z scores for different cognitive domains and total Z-score and global graph theoretical measurements. Fig. 6C shows the significant linear relationship between neuropsychological test scores and graph theoretical measurements. The correlation coefficients are range from 0.19 to 0.25 with uncorrected p < .05.

## 3. Discussion

In this study, we used rMFM whole-brain dynamic modeling to investigate the local and global brain dynamic changes associated with HIV infection. Our results indicate that HIV infection disrupts microscale brain dynamics in several cortical regions, and HIV antiretroviral treatment improves these brain dynamics.

### 3.1. rMFM revealed disruption of local dynamics changes due to HIV infection

Among the areas found to have changes in both recurrent connection strength w and subcortical inputs I were the frontal poles, left lingual gyrus, and right lateral occipital areas. The frontal pole regions are part of the prefrontal cortex thus involved in working memory and multitasking(Gilbert et al., 2006). Disruption in these networks has significant behavioral consequences as shown by studies in HIV-infected subjects with active cocaine use disorder(Meade, Lowen, MacLean, Key, & Lukas, 2011). Our study is also consistent with findings that HIV-infection disrupt the frontostriatal circuitry(Heaton et al., 1995). The lingual gyrus (part of the visual processing system) has also been previously reported to be affected in perinatally HIV-infected youths (Sarma et al., 2019). Ances et al. (Ances et al., 2009; Ances et al., 2010) reported reduced activity and resting cerebral blood flow in the visual cortices during visual stimulation and at rest in HIV-infected subjects using arterial spin labeling(ASL) and fMRI. Wiesman et al.(Wiesman et al., 2018) also demonstrated that the spontaneous and neural oscillatory activity in the visual cortices were affected by HIV-infection using magnetoencephalography (MEG). Our results are complementary to these observations, suggesting that the microscale cortical dynamics including recurrent connection strengths w and subcortical input strength I are affected in these areas in HIV infected individuals.

A number of regions, including left opercular part of inferior frontal gyrus (pars opercularis), superior temporal gyrus, postcentral gyrus, pericalcarine cortex, right insula, and parahippocampal gyrus showed significant differences only in recurrent connections strength w. Some of these, such as the insula and parahippocampal gyrus have been shown to have reduced regional volume and functional connectivity in the HIV-infected population (Kallianpur et al., 2012; McIntosh et al., 2017; Samboju, Philippi, Chan, Cobigo, Fletcher, Robb, Hellmuth, Benjapornpong, Dumrongpisutikul, Pothisri, Paul, Ananworanich, Spudich, Valcour, RV254, et al., 2018). The balanced integration of excitatory and inhibitory synaptic currents in the local cortex allows the cortical network to operate at high efficiency of information transmission at a low energy cost (Yu, Shen, Wang, & Yu, 2018). Although the rMFM model did not disentangle the contribution of excitatory and inhibitory effect, using rMFM method made it possible to compare local recurrent connection changes between groups.

Among the regions that showed significant differences only in subcortical inputs I were the left temporal pole, right temporal pole, right lingual gyrus, right isthmus cingulate, right precuneus, and right superior parietal regions. Some of these regions, such as temporal, right precuneus, superior parietal regions, have been reported to have altered in FC and amplitude of low frequency fluctuations (ALFF) in HIV infected subjects(Yadav et al., 2018). The medial frontal regions (bilateral frontal poles), precuneus and isthmus cingulate gyrus are considered nodes within the default mode network (DMN). HIV-associated decreases in FC within DMN are consistent with previously reports using ICA-based FC analysis (du Plessis et al., 2017; Zhuang et al., 2017).

The bilateral supramarginal gyrus and inferior parietal lobule are considered as the two lateral hubs in the DMN. We found that the recurrent connection strength w of supramarginal gyrus was stronger in the HIV+BSL compared to HC, while the subcortical input strength I in the inferior parietal lobule was less in the HIV+BSL cohort compared to HC cohort (see Supplementary Fig 4.). The significance of increased or decreased w and I are dependent on the local circuit. They are both average inputs that need to be compared to HC. The DMN appears to be a critical network that is sensitive to HIV-associated CNS injury.

In our study, we have shown that after treatment, the microscale local brain dynamics improved in both recurrent connection strength w and subcortical inputs I. Here, change with treatment helps to understand the meaning of the direction in the change observed. Specifically, we found nine out of ten regions’ rMFM metrics in HIV-infected subjects change towards HC levels (Fig. 2. and Fig. 3.). A similar approach was reported by Saenger et al (Saenger et al., 2017) in the context of deep brain stimulation (DBS) in Parkinson’s disease, but to the best of our knowledge our study is the first to use rMFM metrics to assess response in brain networks after cART.

### 3.2. Global and local topological organization disrupted in HIV-infected subjects

Given that rMFM and simulated FC are population-based analyses, we could not directly correlate individual subject cognitive scores and rMFM metrics w and I. We used an intermediate step by applying graph theoretical analysis to empirical FC and then correlated graph metrics to rMFM metrics. We found significant topological changes in FC when comparing HIV+BSL to HC, but no significant change in SC. The local topological measurements showed some association with the cognitive scores. Furthermore, we found after 12-week cART treatment, the functional network topology in HIV-infected subjects also shows transitions towards HC, similar to what was observed in local brain dynamics. Specifically, we observed that smallworldness, global clustering coefficient, and network efficiency were lower in the HIV+BSL (see Fig. 4), which indicated the HIV-infection changes the functional network integration and segregation. A previous study has shown that global clustering coefficient declines with HIV infection (Abidin, D’Souza, Nagarajan, & Wismuller, 2016). The loss of the smallworld organization in brain functional networks has been previously demonstrated in several neurological disorders, such as Alzheimer’s Disease(AD) (Sanz-Arigita et al., 2010), Parkinson’s disease (Baggio et al., 2014), and traumatic brain injury(TBI) (Pandit et al., 2013). In our study, we found that reduced smallworldness in HIV+BSL was associated with decreased neuropsychological composite Z-score, executive function and learning Z-score (Fig. 6C).

The global network efficiency was also reduced in HIV+BSL, suggesting that the communication between different brain regions (van den Heuvel, Stam, Kahn, & Hulshoff Pol, 2009) was affected in HIV-infected subjects. The global network efficiency was associated with the composite Z-score, verbal, executive, and learning Z-score (Fig. 6C). The positive correlation of the network efficiency and executive function performance has been reported in HIV-infected individuals (Ventura et al., 2018), where they focused on the posterior cingulate cortex (PCC).

We and others did not find significant topology difference in structural network of HIV infected individuals (Samboju, Philippi, Chan, Cobigo, Fletcher, Robb, Hellmuth, Benjapornpong, Dumrongpisutikul, Pothisri, Paul, Ananworanich, Spudich, Valcour, Search, et al., 2018; Zhuang et al., 2017). However, two studies (Baker et al., 2017; Bell et al., 2018a) have shown disrupted structural networks in HIV-infected subjects compared to HC in terms of reduced global clustering coefficient, global network efficiency, and connection strength. These two studies differ from our study in the population enrolled. Our patients had been recently diagnosed and had a relatively high CD4 count at baseline (mean 515.8 cells/mm^3^) compared to the other two studies. It is likely that persistence of HIV infection overtime causes some irreversible structural CNS changes (Zhu et al., 2013).

The local topological measurements showed some association with the local dynamic properties, specifically, we found the local topological measurements of FC were also correlated with the recurrent connection strengths w and subcortical inputs I (Fig. 5 and Supplementary Fig. 13). The positive correlations between recurrent connection strength w and clustering coefficient, and local efficiency, respectively, were only found in HC group, suggesting that HIV-infection altered microscale local dynamics, which was also reflected in the change of network topology.

We found the association between microscale local dynamics and empirical network topology using both the Desikan atlas and Destrieux atlas. The HC group consistently shows higher associations than HIV+BSL and HIV+12wk (Fig. 5A, 6B, and Supplementary Fig. 13A, 13B). In HIV+12wk, a significant correlation was only present in Destrieux atlas. This may be due to the inclusion of more cortical parcels in the Destrieux atlas. The results suggest that the cART treatment tends to improve microscale brain dynamics and the efficiency of the brain information transmission.

### 3.3. Limitations

There are some limitations of our approach due in part to the available dataset. First, the rMFM modeling requires reliable and accurate SC reconstruction. Our diffusion data has only one shell at b-value = 1000 s/mm^2^. This single shell scheme may introduce errors for tractography due to fiber-crossing issue (Sotiropoulos et al., 2013). In our study, we have adopted a reliable tractography and SC reconstruction pipeline (Zhang et al., 2018) to mitigate to some extent. Second, we could only derive a population-based whole-brain dynamic model using averaged FC and SC. The reliability of empirical FC is crucial to the rMFM simulation. A recent study from Patricio group (Donnelly-Kehoe et al., 2019) suggests that in order to derive reliable subject-specific brain dynamics, the total length of rsfMRI acquisition should be 20 min with TR=2s. They used both static FC matrices (Pearson correlation of BOLD signal) and dynamic-FC (FCD) metrics to evaluate the simulation results, while in our analysis we only used static FC matrices. Though the FCD estimation requires longer rsfMRI scanning time compared to static FC, our 5 min rsfMRI scan may still limit the static FC reliability to generate subject-specific FC. In this regard, we attempted to derive subject-specific rMFM models using individual SC and FC maps, however the variance of the similarities was much greater than that of the population-based model. We thought this large inter-subject variation for rMFM came from both individual differences and SC/FC noise. It is difficult to disentangle these two effects in our current data. However, we have shown that using group-averaged population-based rMFM modeling is much more stable across different cohorts, and we validated these findings using a different atlas. Thus, the rMFM model parameters (recurrent connection strength w, and subcortical input strength I) and the simulated FC can only be used in comparing groups but not individual parameters, such as CD4 count or cognitive scores. Lastly, our sample size is relatively small, constraining the stability of rMFM modeling as we needed to split the cohorts into training and test groups. Unfortunately, enrolling cART naïve patients is quite challenging. Future studies using more advanced diffusion scheme and longer rsfMRI acquisition, should provide more reliable subject-specific rMFM model reconstruction.

### 3.4. Conclusion

We investigated the effect of HIV infection on the microscale local dynamics derived by whole-brain computational modeling. We have identified several brain regions where recurrent connection strengths and subcortical inputs differ among HC, HIV+ untreated and HIV+ after treatment. Treatment improved local brain dynamics, and this was also reflected in improved brain network topology and cognitive performance. However, short-term treatment did not fully reverse CNS injury. Whether further diverge on CNS injury and cognitive performance occurs between HIV+ and HIV-over longer periods of time will need to be addressed in future studies. In this regard, whole-brain dynamic modeling is a promising approach for assessing CNS injury progression and response to interventions.

## 4. Materials and Methods

### 4.1. Ethics statement

The study was reviewed and approved by the Institutional Review Board at the University of Rochester Medical Center and all subjects signed a written informed consent prior to undergoing study procedures. All subjects underwent a comprehensive clinical, laboratory (chemistry, hematology, and urine analysis), neurocognitive, and neuroimaging evaluation. HIV-infected individuals were assessed before and 12 weeks after starting cART, while HIV-uninfected controls were assessed only once at baseline. Subjects’ demographics are listed in Table 1 in results session.

**Table 1.** Demographics and baseline clinical variables

|  | HIV Infected<br>(n=42) | Healthy Control<br>(n=46) | p-value |
| --- | --- | --- | --- |
| Age (years) Mean±SE | 34.9±2.2 | 37.3±2.1 | 0.196 |
| Gender (M:F) | 39:3 | 23:23 | <0.001 |
| Ethnicity<br>(White:Black:Other) | 22:19:1 | 35:5:6 | 0.042 |
| Education |  |  | 1 |
| ≤12 years | 12 | 6 |  |
| >12 years | 30 | 40 |  |
| HIV duration by patient<br>report at baseline (months)<br>Median (Lower Quantile,<br>Upper Quantile) | 1(1, 12.75) | NA | -- |
| Baseline CD4 cell count<br>(cells/mm<sup>3</sup>) Mean±SE | 515.8±42.3 | NA | -- |
| Baseline HIV RNA levels<br>(log<sub>10</sub> unit) Median (range) | 4.5 (1.7, 5.8) | NA | -- |
| HAND Classification at<br>Baseline |  |  | 1 |
| Normal | 21 | NA |  |
| ANI | 20 | NA |  |
| MND | 1 | NA |  |
| Total NPZ Score Mean (SD) |  |  | 0.014 |
| BSL | -1.886(3.79) | 0.100(3.94) |  |
| 12-week | -0.187(3.88) | -- |  |

HAND= HIV-Associated Neurocognitive Disorder; ANI= Asymptomatic Neurocognitive Impairment; MND= Mild Neurocognitive Disorder; BSL = HIV+ and HC at Baseline; 12-week = HIV+ after 12-week cART treatment.

### 4.2. MRI data collection

MRI was performed on a 3T Siemens MAGNETOM Trio MRI scanner (Siemens Healthineer, Germany) equipped with a 32-channel head coil. T1-weighted three-dimensional magnetization-prepared rapid acquisition gradient echo (MPRAGE) images were acquired, with repetition time (TR)/inversion time (TI)/echo time (TE) = 2530/1100/3.44 ms, voxel size = 1.0 × 1.0 × 1.0 mm^3^, flip angle = 7°, bandwidth = 190 Hz/pixel. Diffusion weighted imaging (DWI) data was acquired with the following parameters: TR/TE = 34.85/7.12 ms; 10 b = 0 s/mm^2^ images; 60 diffusion weighting images with b = 1000 s/mm^2^ and direction uniformly distributed on a unit sphere; voxel size = 2 × 2 × 2 mm^3^, bandwidth = 1502 Hz/pixel. A double-echo gradient echo field map sequence was acquired with the same resolution as the DWI sequence and was used to correct for distortion caused by B0 inhomogeneity. Resting-state fMRI data was acquired using a gradient echo-planar imaging (EPI) sequence, with TR/TE = 2000/30 ms, voxel size = 4 × 4 × 4 mm^3^, 150 time points, flip angle = 90°, bandwidth = 1562 Hz/pixel. During the entire 5-min resting-state fMRI series, participants were instructed to keep their eyes closed and avoid falling asleep.

### 4.3. Overview of data processing

The processing pipeline is shown as Fig. 7.

1. We preprocessed the T1-weighted (T1w) image, DWI, and rsfMRI, and constructed the empirical SC and FC (Section 4.4).
2. The rMFM whole-brain model was built for each group (Section 4.5): We split the empirical SC and FC into training and test datasets for each cohort. We then derived the rMFM model parameters using the training dataset and validated each rMFM model using the test datasets (Section 4.6). The microscale brain properties, namely recurrent connection strength, denoted as w, and subcortical input strength, denoted as I, were derived from each group were then compared among HIV+BSL, HIV+12wk and HC subjects.
3. Graph topological analysis on empirical FC and SC (Section 4.7): We investigated the topological changes in empirical FC and correlated the recurrent connection strength w, and subcortical inputs I with the nodal graph theoretical measurements.
4. The rMFM modeling and graph theoretical analysis were validated in three steps (see supplementary material): a) split the data into different training and testing groups, b) doubled the rMFM optimization steps, c) reproduced our results using a finer segmented brain atlas, Destrieux atlas (Destrieux, Fischl, Dale, & Halgren, 2010), which includes 148 cortical regions.
5. Neuropsychological assessment scores were compared among cohorts, and their relations with graph theoretical measurements were also investigated (Section 4.8).

**Fig. 7.**
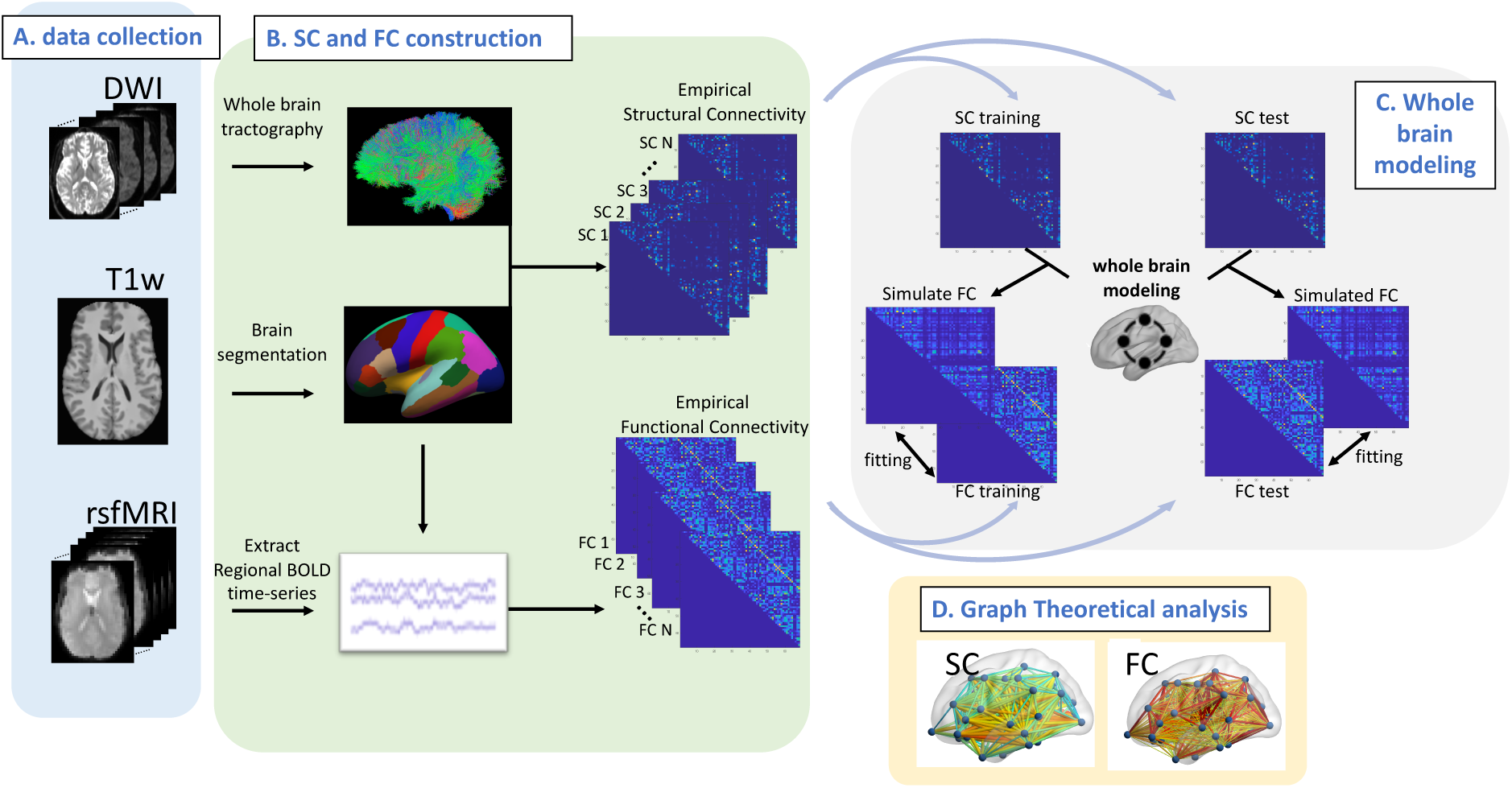
Overview of processing pipeline. A) We collected diffusion weighted image (DWI), T1-weighted (T1w) structural image, and resting-state functional MRI (rsfMRI) for each HIV+ and HC subject. B) For each subject, the T1w image was segmented using Freesurfer. DWI and rsfMRI were preprocessed and co-registered to T1w image. Whole-brain tractography for each subject was derived from DWI image, and a structural connectivity (SC) matrix was generated for each subject. The streamline count for each pair of brain regions constitutes the SC matrix. Regional BOLD time series were extracted for each subject. The Pearson’s correlation coefficient of the BOLD time series of two brain regions constitutes the FC matrix. C) Whole-brain dynamic modeling, specifically, relaxed mean field dynamic modeling (rMFM), is used to study the neuronal dynamic changes between HIV+ subjects and HC subjects. D) We also used graph theoretical analysis to investigate the topological changes by comparing the SC and FC for HIV+ subjects with HC subjects.

### 4.4. MRI data preprocessing

#### 4.4.1. Anatomical T1w image analysis

Three-dimension high resolution T1w images were brain extracted using Brain Extraction Tool (Smith, 2002) (BET) to remove nonbrain tissue. Cortical and subcortical segmentation were then performed with Freesurfer image analysis suite (http://surfer.nmr.mgh.harvard.edu/). The Desikan-Killiany atlas (Desikan et al., 2006) and the Destrieux atlas (Destrieux et al., 2010) were mapped to individual T1w images and were further used for SC and FC construction. The 68 labeled cortical regions, 34 in each hemisphere from the Desikan-Killiany atlas are served as nodes in the SC and FC. The Destrieux atlas includes 148 labeled cortical regions, 74 in each hemisphere. The main results in the following sections are calculated using Desikan-Killiany atlas, whereas the Destrieux atlas was used for validation of rMFM modeling results (supplementary materials).

#### 4.4.2. DTI data processing and whole-brain tractography

Preprocessing of DTI data was reported in detail previously (Zhuang et al., 2017). Briefly, the b0 images and diffusion-weighted images were motion corrected using a six-degrees of freedom (DOF) rigid-body registration and field maps were used to correct the susceptibility induced distortion, using FUGUE in FSL (Jenkinson, 2003). DTI processing and structural connectome construction were then performed using the population-based structural connectome pipeline (PSC) (Zhang et al., 2018). A reproducible probabilistic whole-brain tractography algorithm (Girard, Whittingstall, Deriche, & Descoteaux, 2014; Maier-Hein et al., 2017) was used to reconstruct streamlines. The tissue partial volume estimation (PVE) maps obtained from the anatomical T1w image helped reduce the tractography bias. The particle filtering tractography (PFT) was used to reconstruct more reliable streamlines (Girard et al., 2014). After whole-brain tractography was generated, the Desikan-Killiany atlas and Destrieux atlas were used to construct the structural connectome represented by 68×68 matrix and 148×148 matrix, respectively. The individual SC matrix was used for graphic theoretical analysis. Subjects were randomly split into training and test groups, controlling for age, so that there was no significant age difference between the training and testing group, and between the three cohorts (see Supplementary Table 1). For the whole-brain dynamic modeling, the SC matrices were averaged across subjects in training and test datasets, respectively.

#### 4.4.3. Resting-state functional MRI analysis

Resting-state functional MRI data was pre-processed using FEAT (FMRI Expert Analysis Tool) (Woolrich, Ripley, Brady, & Smith, 2001), part of FSL(Jenkinson, Beckmann, Behrens, Woolrich, & Smith, 2012). Detailed preprocessed steps are included in the supplementary materials. The pre-processed fMRI data was then further cleaned by using FIX (Griffanti et al., 2014; Salimi-Khorshidi et al., 2014), an ICA-based noise removal software that automatically classifies noise components of the resting-state functional data. Head motion, white matter, and cerebral spinal fluid (CSF) time series were regressed out as nuisance regressors. Two HIV+BSL subjects and one HC subject were removed from rsfMRI analysis due to head motion greater than 2.5mm. We used a two-sample t-test to ensure there was no significant head motion difference between HIV+ and HC cohort (HIV+ mean displacement = 0.748mm, HC mean displacement = 0.645mm, p = 0.065).

Mean BOLD signals were then extracted from each parcel in the Desikan-Killiany and Destrieux atlases. Pearson’s correlation of the mean BOLD signals between each pair of parcels was calculated yielding an FC matrix for each subject. The FC matrix were then Fisher’s r-to-z transformed, resulting in a zFC matrix. The zFC matrices for all subjects were used for graph theoretical analysis. In preparation for rMFM whole-brain dynamic modeling, the zFC matrices were averaged across subjects within training or test dataset, for each age-controlled cohort (HIV+BSL, HIV+12wk, and HC) respectively (Supplementary Table 1).

### 4.5. Simulated FC using rMFM model

The whole-brain dynamic mean-field model (MFM) (Deco et al., 2014; Deco et al., 2013) has been widely used to derive spontaneous brain activity from structural connectivity (Deco et al., 2018). Here we used a modified version of MFM, the rMFM (P. Wang et al., 2019), which assumes the recurrent connection strengths w and subcortical inputs I are not uniformly distributed in the brain. This model has been previously proved to improve the FC simulation by 53% over the original MFM (P. Wang et al., 2019). Using rMFM, we simulated the neural activity for each cortical region, and then used the Balloon-Windkessel hemodynamic model (Friston, Harrison, & Penny, 2003; Friston, Mechelli, Turner, & Price, 2000) to convert neural activity to simulated BOLD signal. The simulated FC was then calculated using Pearson’s correlation of the BOLD signal for pairwise cortical regions. More details of the rMFM model and its mathematical relations to various parameters are given in supplementary materials.

### 4.6. Model parameters estimation

Empirical SC and FC for each group (HIV+BSL, HIV+12wk, and HC) in the training dataset was used to estimate the model parameters. There are 138 parameters to be optimized when we use the Desikan atlas (68 recurrent connection strength w_i_, 68 subcortical input strength I_i_, global scaling factor G, and noise coefficient *σ*).

The optimization steps were based on the expectation-maximization algorithm in dynamic causal modeling (DCM) (Friston et al., 2003), and were detailed in (P. Wang et al., 2019). In this study, 500 iterations were performed for each run, and the final optimum estimated parameters for the rMFM model were chosen from the highest Pearson’s correlation coefficient between the empirical FC and simulated FC in the training dataset from the 500 iterations (highest similarity for each cohort in Supplementary Fig. 1.A). The corresponding 138 model parameters were stored, shown as one column in Supplementary Fig. 2.

We noticed that the random initialization parameters had a small effect on the final model parameters, so we repeated the entire optimization process with 25 different random initializations for each cohort. We then chose the top 5 sets of model parameters, i.e. the model parameters corresponding to the 5 highest Pearson’s correlation coefficients between empirical FC and simulated FC (the five highest similarities in Supplementary Fig. 1.B).

We then fed the empirical SC in the test dataset for each group to its own rMFM model, to validate these three rMFM models by calculating the similarity between the simulated FC and empirical FC for each group. After each model for HIV+BSL, for HIV+12wk, and for HC was validated, we obtained three rMFM generative models representing the three cohorts.

The recurrent connection w_1_ = (w_1_,…, w_n_} and subcortical inputs I = I_1_,…, I_n_}, where n = 68 for Desikan-Killiany atlas, in each rMFM model represent the microscale brain dynamics for each group and were compared among groups.

To ensure 500 iterations for each optimization run were sufficient, we also repeated the process with 1000 iterations, with 15 different random initializations for each group. In addition, we repeated the rMFM modeling process using a different atlas to validate our results. To do this, we constructed the empirical SC and FC using Destrieux atlas, which yields 298 model parameters to be optimized. Detailed results from these analyses are reported in the supplementary materials.

### 4.7. Graph theoretical analysis

The topology of the empirical FC and SC was further evaluated using graph theoretical measurements (Rubinov & Sporns, 2010), with calculations using Brain Connectivity Toolbox (BCT, https://sites.google.com/site/bctnet/) and graph theoretical network analysis toolbox (GRETNA (J. Wang et al., 2015), https://www.nitrc.org/projects/gretna).

It is known that there are some spurious connections in the connectivity matrix that should be taken into consideration in the analysis. Also, applying arbitrary thresholding of the FC or SC metrics before calculating the topology properties may influence the result (van Wijk, Stam, & Daffertshofer, 2010). Therefore, we applied a range of thresholds to study the topology properties under different network sparsity. The graph theoretical measurements were calculated over 10 different network sparsity values ranging from 0.05–0.5, where the network sparsity is defined as the ratio of the number of edges divided by the maximum possible number of edges in a network. Undirected weighted matrices were used in these calculations.

Four common topological measurements were chosen in this study for analysis of network properties, including the clustering coefficient, shortest path length, global efficiency, and smallworldness. Detailed information is included in supplementary materials. We evaluated the association between the nodal graph theoretical measurements and the recurrent connection strength w, and the subcortical inputs I, using Pearson’s correlation.

### 4.8. Neuropsychological assessment

The neurocognitive evaluation was performed by trained staff and supervised by a neuropsychologist, and included tests of executive function (Trailmaking Test Part B, Stroop Interference Task), speed of information processing (Symbol Digit Modalities Test and Stroop Color Naming), attention and working memory (CalCAP(CRT4) and WAIS-III Letter-Number Sequencing), learning (Rey Auditory Verbal Learning Test RAVLT (trials 1–5), Rey Complex Figure Test Immediate Recall), memory (Rey Auditory Verbal Learning Test RAVLT Delayed Recall, Rey Complex Figure Test Delayed Recall), verbal fluency (Letter, Category and Action Fluency Tasks), and motor (Grooved Pegboard, the left and right hands). An estimate of premorbid intellectual functioning ability was obtained via WRAT-4 Reading. The total composite Z-score was the primary cognitive outcome and was created from the linear combination of the Z-scores of the seven cognitive domains measured (executive function, speed of information processing, attention and working memory, learning, memory, verbal fluency and motor). HIV-Associated Neurocognitive Disorder (HAND) diagnoses were determined for each participant according to the Frascati criteria(Antinori et al., 2007). The neuropsychological composite Z-score and the seven cognitive domains Z-scores were compared between HIV+BSL, HIV+12wk, and HC respectively. We also investigated the relationship between the graph theoretical measurements with cognitive performance scores.

### 4.9. Statistical Analysis

Comparisons of continuous variables between two independent groups were conducted by two-group Welch’s unequal variances t-test. Fisher’s exact test was used to test any proportional differences in race, gender, and education level between the HIV+ and Control groups in Table 1. Pearson’s correlation test was used to analyze the association between two continuous variables. A p-value p < 0.05 was considered statistically significant for a single hypothesis testing problem. For inferential problems that involved multiple hypotheses, Benjamini–Hochberg multiple testing procedure was used to control the false discovery rate (FDR) at < 0.05 level. The statistical analysis in this study was performed using MATLAB 2017b (https://www.mathworks.com/products/matlab.html) and R version 3.6.1 (https://www.r-project.org/).

## 4.10. Data Availability

The datasets from the current study are available from the corresponding author on reasonable request. The codes are available at https://github.com/yzhuang4/whole_brain_modeling. The codes for rMFM model used in the paper are adopted from https://github.com/ThomasYeoLab/CBIG/tree/master/stable_projects/fMRI_dynamics/Wang2018_MFMem.

## Acknowledgements

We thank Alicia Tyrell for providing data management; Arun Venkataraman, Kyle Murray for the insightful discussion and comments during preparation of this paper.

